# Recovery of monocyte exhaustion is associated with resolution of lung injury in COVID-19 convalescence

**DOI:** 10.1101/2020.10.10.20207449

**Authors:** N.A. Scott, S.B. Knight, L. Pearmain, O. Brand, D.J. Morgan, C. Jagger, S. Khan, P. Hackney, L. Smith, M. Menon, J. E. Konkel, H. A. Shuwa, M. Franklin, V. Kästele, S. Harbach, S. Brij, A. Ustianowski, A. Uriel, G. Lindergard, N. Diar Bakerly, P. Dark, A.G. Mathioudakis, K.J. Gray, G.M. Lord, T. Felton, C. Brightling, L-P Ho, NIHR Respiratory TRC, CIRCO, K. Piper Hanley, A. Simpson, J.R. Grainger, T. Hussell, E.R. Mann

## Abstract

Severe acute respiratory syndrome coronavirus 2 (SARS-CoV-2) infection resulting in the clinical syndrome COVID-19 is associated with an exaggerated immune response and monocyte infiltrates in the lungs and other peripheral tissues. It is now increasingly recognised that chronic morbidity persists in some patients. We recently demonstrated profound alterations of monocytes in hospitalised COVID-19 patients. It is currently unclear whether these abnormalities resolve or progress following patient discharge. We show here that blood monocytes in convalescent patients at their 12 week follow up, have a greater propensity to produce pro-inflammatory cytokines TNFα and IL-6, which was consistently higher in patients with resolution of lung injury as indicated by a normal chest X-ray and no shortness of breath (a key symptom of lung injury). Furthermore, monocytes from convalescent patients also displayed enhanced levels of molecules involved in leucocyte migration, including chemokine receptor CXCR6, adhesion molecule CD31/PECAM and integrins VLA-4 and LFA-1. Expression of migration molecules on monocytes was also consistently higher in convalescent patients with a normal chest X-ray. These data suggest persistent changes in innate immune function following recovery from COVID-19 and indicate that immune modulating therapies targeting monocytes and leucocyte migration may be useful in recovering COVID-19 patients with persistent symptoms.

## Main

The Coronavirus Immune Response and Clinical Outcomes (CIRCO) study was carried out at four hospitals in Greater Manchester, UK, and was designed to examine immune responses in COVID-19 patients during primary admission (acute phase) and recovery (convalescence; **Figure 1**). We recently demonstrated profound alterations in the monocytes of COVID-19 patients which correlated with disease severity and could potentially be used to identify patients destined for a severe disease outcome upon admission^1^. Immune dysfunction in monocytes was a defining feature of patients with severe COVID-19 disease. Whether monocyte abnormalities persist during convalescence following patient discharge is currently unknown. Here, we demonstrate distinct alterations in monocytes in convalescent COVID-19 patients during recovery, which correlate with radiographic features of disease and shortness of breath (dyspnoea).

**Figure 1.**
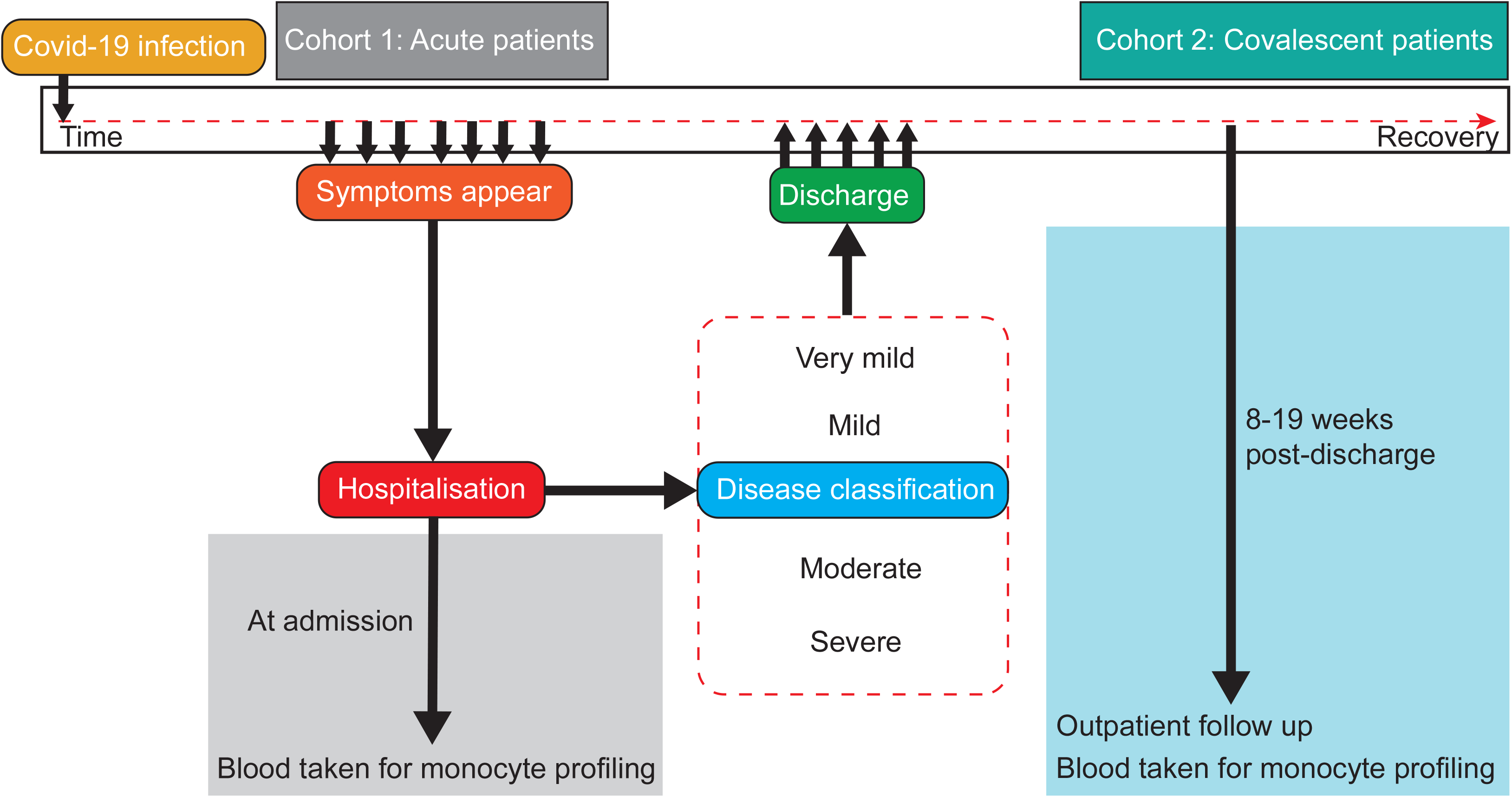
Patient recruitment and categorization. Acute patients were recruited to the study as close to admission as possible, and covalescent patients were recruited during outpatient follow up appointment 8-19 weeks after hospital discharge. Peripheral blood samples were collected on recruitment and samples were analysed immediately.

Between 14^th^ June and 25^th^ August 2020, 44 patients attending outpatient follow up at median of 87 days (IQR 79.5-99 days) after discharge following inpatient admission for COVID-19 were included as “convalescent” patients. COVID-19 cases were defined by a positive RT-PCR test for SARS-CoV-2 or characteristic symptoms and radiographic changes of COVID-19 in the absence of a positive test. Additionally, patients were included where full clinical and monocyte data were available. One patient was excluded due to a confounding non-COVID acute illness at the time of COVID-19 admission. All of the convalescent patients had abnormal chest X-rays at the time of primary hospital admission and were retrospectively scored for disease severity according to the degree of respiratory failure **(Extended data 1**). At outpatient follow up, 26% of convalescent patients continued to display abnormalities on chest imaging attributed to COVID-19. 36% of convalescent patients with no chest imaging abnormalities remained breathless compared with 64% of patients who continued to exhibit an abnormal chest X-ray (**Figure 2)**.

**Figure 2.**
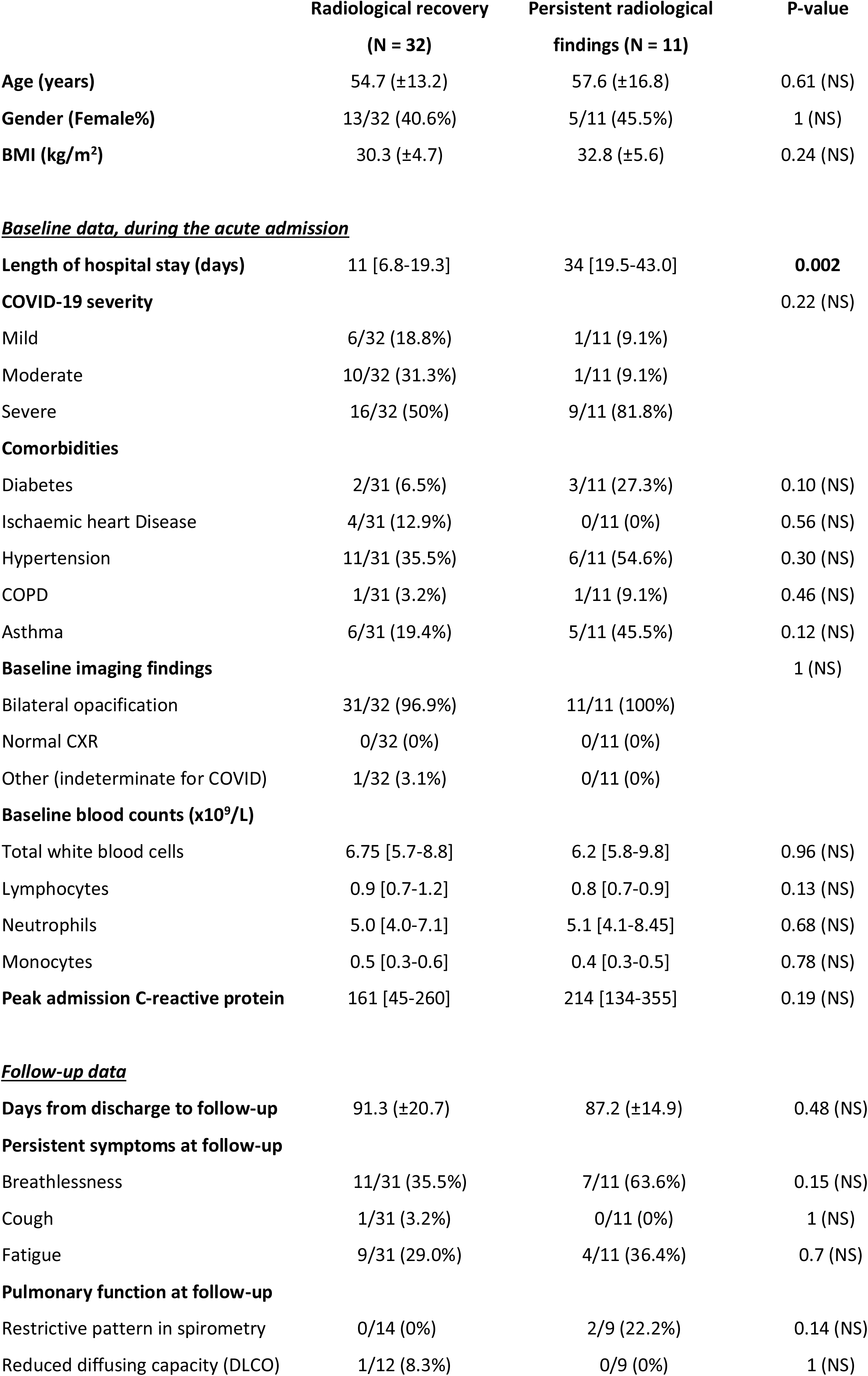
Clinical characteristics of convalescent patients. Normally distributed continuous data presented as mean (± standard deviation) and compared using t-test; while the remaining continuous data are presented as median [interquartile range] and compared using Mann-Whitney U test. Dichotomous data are presented as percentages and compared using chi-squared or the Fisher test, as appropriate.

Two comparator groups were included in analysis; twenty “acute” patients (in-patients with COVID-19) were recruited during the same time period as convalescent COVID-19 patients, and healthy controls (**Extended data 2)**. Three patients were excluded due to concurrent confounding acute illnesses.

Monocytes can contribute significantly to inflammatory disease directly or via their differentiation into macrophages or dendritic cells in peripheral tissues; recent reports demonstrate infiltrates of monocytes in the lungs, kidney, heart, spleen, and muscle from post-mortem tissue of deceased COVID-19 patients^2-4^. Failure of lung injury resolution often leads to the development of pulmonary fibrosis, in which monocytes and macrophages play a key role^5^, and is a complication of great concern following COVID-19 infection. We, and others, have previously demonstrated that during acute infection and hospitalisation, monocytes from COVID-19 patients exhibit a restricted capacity to produce inflammatory cytokines in response to activation with microbial stimuli^1,6^. The reasons for this are unclear, but may signify exhaustion of immune cells in COVID-19 patients, which is supported by the rapid reduction in systemic levels of inflammatory cytokines following admission to intensive care^1^.

The restoration of monocyte responses in recovery from COVID-19 has not been addressed to date. We now show that following stratification of convalescent patients at follow up, production of the inflammatory cytokines IL-6 and TNFα by monocytes in response to stimulation with lipopolysaccharide (LPS) was consistently higher in convalescent patients with normal chest X-rays and no dyspnoea (**Figure 3a**). To determine whether enhanced cytokine production by monocytes was unique to convalescence, we compared convalescent patients to hospitalised acute COVID-19 patients and healthy controls. Monocytes from acute patients produced levels of the inflammatory cytokines IL-6, TNFα, IL-1β and CCL2 comparable to healthy controls, in contrast to their convalescent counterparts that demonstrated increased proportions of monocytes producing IL-6 and TNFα (**Figure 3b**). Production of IL-6 by monocytes was correlated with production of TNFα in convalescence **(Figure 3c)**.

**Figure 3.**
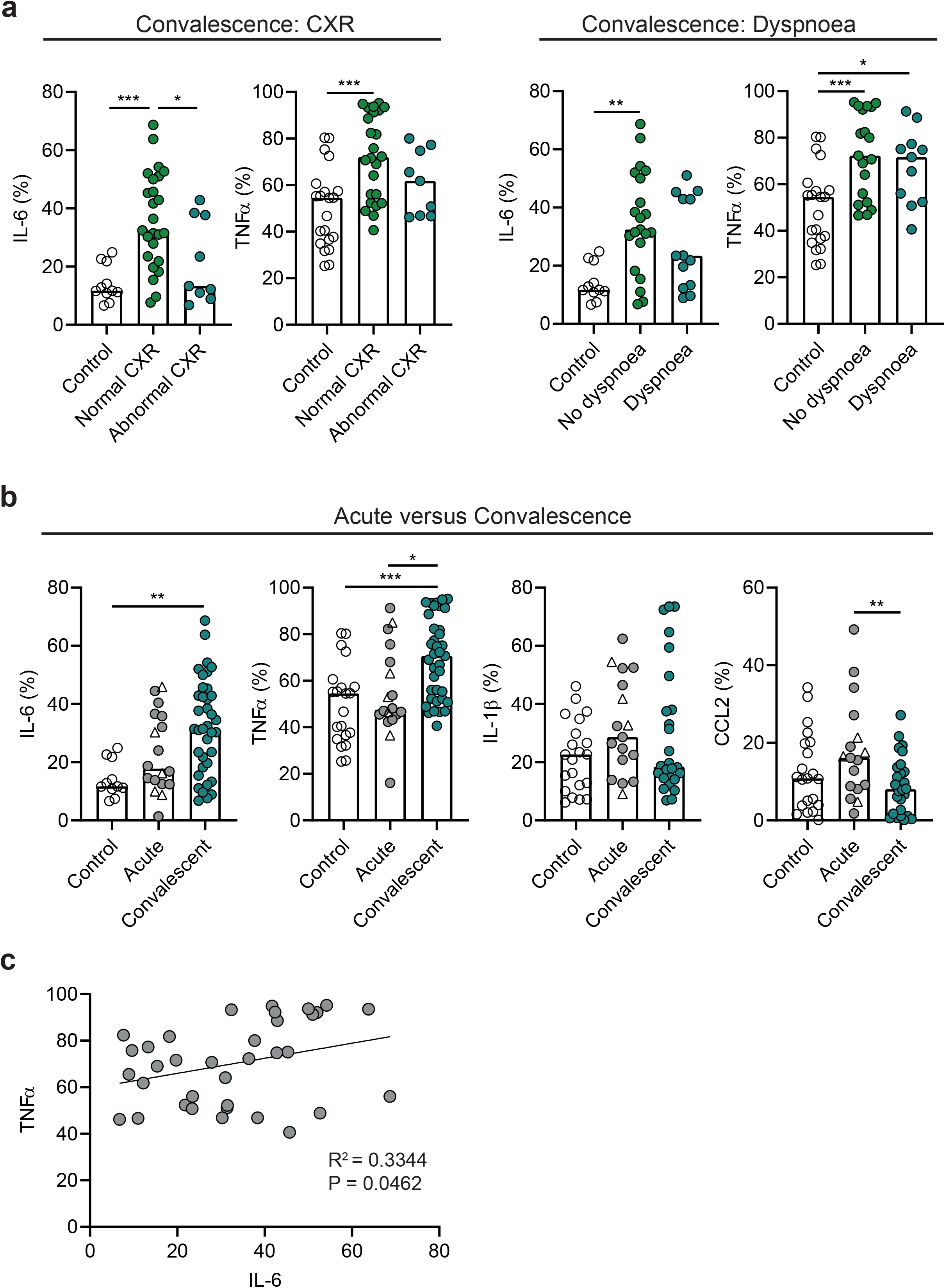
Cytokine production by circulating monocytes in COVID-19 convalescent patients. **(A)** Graphs show proportions of circulating monocytes producing IL-6 and TNF*α* following LPS stimulation of freshly prepared PBMCs from healthy individuals (IL-6: n=11, TNF*α*: n=21), convalescent COVID-19 patients with normal chest X-ray (CXR; IL-6: n=25; TNF*α*:n=22), abnormal CXR (IL-6: n=9; TNF*α*: n=9), dyspnoea (IL-6: n=21; TNF*α*: n=19) or no dyspnoea (IL-6:n=13; TNF*α*: n=11). **(B)** Graphs show proportions of circulating monocytes producing IL-6, TNf*α*, IL-1*?* and CCL-2 following LPS stimulation of freshly prepared PBMCs from healthy individuals (IL-6: n=11, TNF*α*: n=21), acute admission COVID-19 patients (n=18), and convalescent COVID-19 patients (n=37). **(C)** Correlations of stimulated monocytes from convalescent COVID-19 patients producing IL-6 versus those producing TNF*α* (n=35). Within the acute admission group, open triangles represent “very mild” patients which have minimal/no substantial changes on CXR related to COVID-19 during acute admission. One way ANOVA with Holm-Sidak post-hoc test: 3A, B. Spearman ranked coefficient correlation test: 3C. (P*<0.05; **P<0.01; ***P<0.001; ****P<0.0001).

We next examined whether the ability of monocytes from COVID-19 convalescent patients to produce inflammatory cytokines was related to their disease severity during acute admission. Monocytes from those patients who had moderate or severe disease in hospital during acute disease produced more IL-6 (but not TNFα) during convalescence compared to previously mild patients **(Extended data 3a)**. Taken with abnormal chest X-ray and dyspnoea in recovery being predominant in patients with moderate and severe disease during acute phase (**Figure 2)**, the discordance between IL-6 and dyspnoea/abnormal chest X-ray at follow up indicate that recovery of monocyte exhaustion and restoration of cytokine production is important in resolving lung injury following acute SARS-CoV-2 infection. Although IL-6 is implicated in lung injury pathogenesis^7^, blockade of IL-6 can worsen lung injury^8^. This dual role for IL-6 in lung injury is likely dependent on the local cytokine milieu and the timing of the disease course.

Linear regression analysis comparing monocyte cytokine production in convalescence with clinical parameters measured at the time of acute admission including CRP levels, FiO2, monocyte count, neutrophil count or lymphocyte count showed no significant correlations **(Extended data 3b)**. Age or BMI during acute COVID-19 did not affect TNFα or IL-6 production by monocytes in convalescence **(Extended data 3c)**. Although monocytes from male COVID-19 convalescent patients had a propensity to produce more TNFα than their female counterparts, gender did not impact on IL-6 production (**Extended data 3d)**. Furthermore, monocytes from male convalescent patients produced heightened levels of TNFα compared to male acute patients and male controls (**Extended data 3e)**, indicating the heightened production of TNFα in convalescent patients was not due to differences in male:female ratios between groups. Overall, these data indicate that enhanced monocyte responsiveness following SARS-CoV-2 infection correlates with resolution of lung injury in recovery. Interestingly, in mouse models of pulmonary fibrosis, TNFα accelerates resolution of fibrosis^9^ and may perform a similar function in COVID-19 in terms of resolution of lung injury.

Given that enhanced monocytic infiltrates are found in post-mortem specimens of the lungs and peripheral tissues of COVID-19 patients^2-4^, we next examined monocytes in convalescent COVID-19 patients for altered expression of molecules involved in cell migration including chemokine receptors, adhesion molecules and integrins. As with the cytokine data, we examined for correlations between migration markers and current abnormal chest X-ray. Integrins LFA-1 and VLA-4, adhesion molecule CD31/PECAM and chemokine receptor CXCR6 on monocytes in convalescence were consistently higher in patients with fully resolved radiographic features of COVID-19 (normal chest X-rays; **Figure 4a**). A greater proportion of monocytes in acute admission COVID-19 patients also expressed CXCR6 compared to controls as well as in convalescence, but although nearly all monocytes expressed adhesion molecule CD62L in all cases, levels of CD62L expression on monocytes were heightened in the acute admission group only (**Figure 4b**). CD62L levels were normal in the convalescent group (**Figure 4b)**, but several migration molecules were aberrantly expressed on monocytes from convalescent patients only, rather than during active disease, including LFA-1, VLA-4, and CD31/PECAM (**Figure 4c)**.

**Figure 4.**
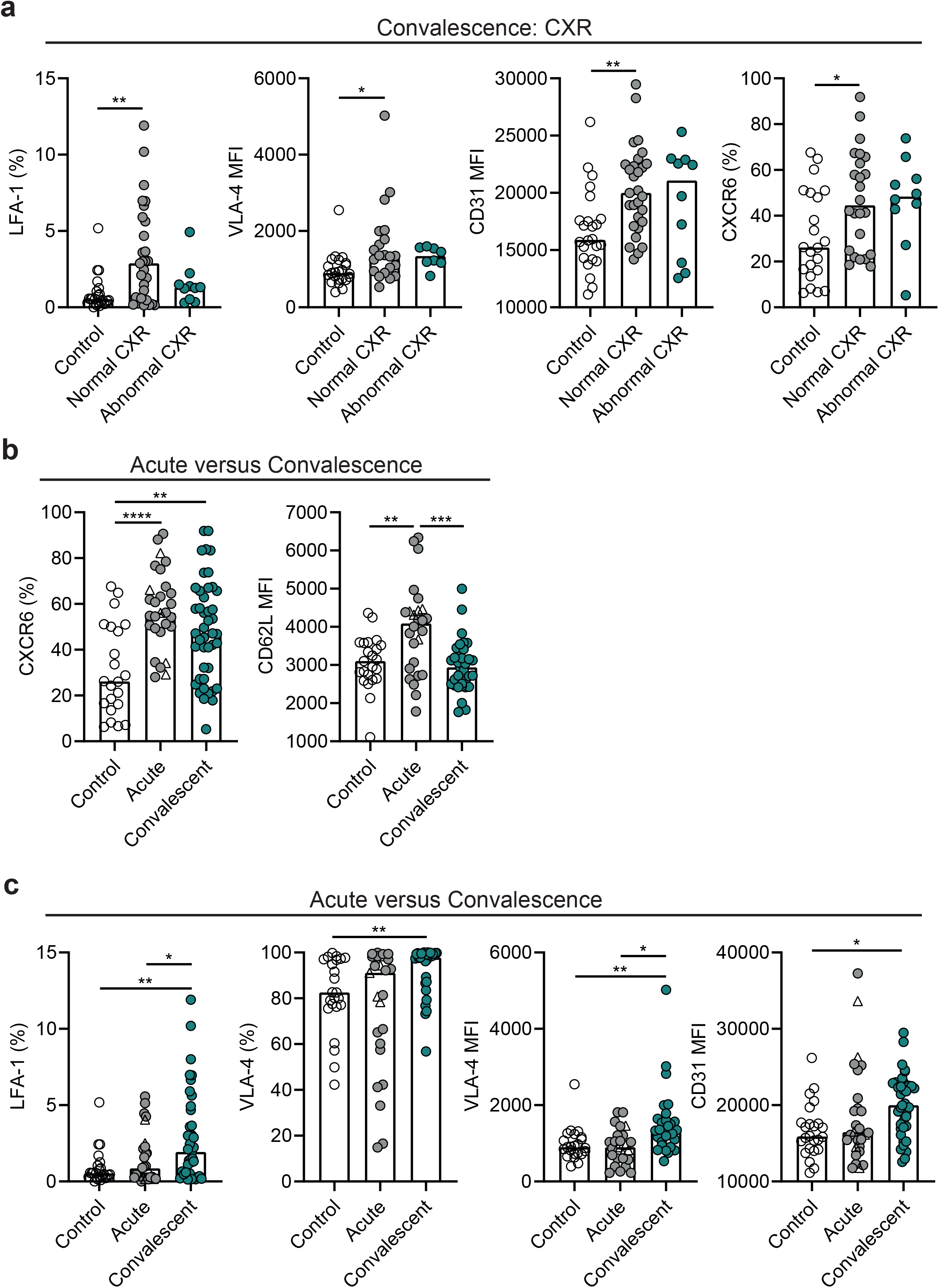
Migration marker expression by circulating monocytes in COVID-19 convalescent patients. **(A)** Graphs show proportions of circulating monocytes expressing LFA-1 and CXCR6 in healthy individuals (LFA-1: n=23; CXCR6: n=22), convalescent patients with normal CXR (LFA-1: n=29; CXCR6: n=24) and abnormal CXR (LFA-1: n=10; CXCR6: n=10), and graphs show levels of expression of VLA-4 and CD31 as assessed by mean fluorescence intensity (MFI) from healthy individuals (VLA-4: n=23; CD31: n=24), convalescent patients with normal CXR (VLA-4: n=21;CD31: n=29) and abnormal CXR (VLA-4: n=8; CD31: n=10). **(B)** Graphs show proportions of circulating monocytes expressing CXCR6 and levels of expression of CD62L as assessed by MFI from healthy individuals (CXCR6: n=22; CD62L: n=23), acute admission COVID-19 patients (CXCR6: n=28; CD62L: n=26) and convalescent COVID-19 patients (CXCR6: n=44; CD62L: n=33). **(C)** Graphs show proportions of circulating monocytes expressing LFA-1 and VLA-4, and levels of expression of VLA-4 and CD31 as assessed by MFI, from healthy individuals (LFA-1: n=23; VLA-4: n=23, VLA-4 MFI: n=23; CD31 MFI: n=24), acute admission COVID-19 patients (LFA-1: n=28; VLA-4: n=25, VLA-4 MFI: n=25; CD31 MFI: n=28) and convalescent COVID-19 patients (LFA-1: n=39; VLA-4: n=29, VLA-4 MFI: n=29; CD31 MFI: n=39). Within the acute admission group, open triangles represent “very mild” patients which have minimal/no substantial changes on CXR related to COVID-19 during acute admission. One way ANOVA with Holm-Sidak post-hoc test: 4A (CD31 MFI and CXCR6), 4B, 4C. Kruskal-Wallis test with Dunn’s post-hoc test: 4A (LFA-1 and VLA-4 MFI). (P*<0.05; **P<0.01; ***P<0.001; ****P<0.0001).

Monocytes utilize CD31/PECAM-1 for migration through the endothelium^10^, in the process of crossing the endothelial lining of blood vessels to enter tissues (transendothelial migration). VLA-4 is used for adhesion of activated endothelium^11^ and LFA-1 for intralumenal crawling^12^. These molecules thereby facilitate monocyte infiltration into peripheral tissues with CXCR6 directing migration towards CXCL16 produced in tissues under inflammatory conditions^13^. Our data indicate that even after the acute disease period, monocytes from COVID-19 patients demonstrated increased potential to migrate into peripheral tissues compared to controls during recovery. CD62L plays a key role in regulating recruitment of monocytes to lymphoid tissue during inflammation^14^; thus the enhanced expression of CD62L on monocytes in acute patients suggests increased migration of monocytes to lymph nodes during active SARS-CoV-2 infection, but is resolved during convalescence. Enhanced migration to lymphoid tissue during infection would also at least in part explain why there are no changes in circulating monocyte numbers in acute patients despite indications of enhanced egress of monocytes from bone marrow during COVID-19 from our previous findings^1^.

Expression of these migration-associated molecules by monocytes in convalescent patients was not correlated to the level of disease severity from acute admission apart from VLA-4 which was only significantly higher in convalescent patients who had severe or moderate COVID-19 upon their primary admission to hospital (**Extended data 4)**. Monocyte expression of migration molecules in convalescent patients was not correlated to clinical parameters measured during previous hospitalisation including CRP levels, Fi02, monocyte count, neutrophil count or lymphocyte count (**Extended data 5)**. Migration molecule expression was not correlated to age, gender or BMI (**Extended data 6)**.

The mechanisms by which monocytes may be involved in resolution of lung injury are unclear, but there are implications for repopulation of tissue resident macrophage populations in the lung that contribute directly to tissue repair. Overall, these data indicate that restored activation of monocytes upon stimulation and alterations in the expression of molecules associated with migration are associated with lung injury resolution following SARS-CoV-2 infection. Reprogramming of innate immune cells after exposure to pathogens has been reported previously. This phenomenon has been associated with epigenetic reprogramming, known as trained immunity, and confers non-specific immunity from secondary infections including respiratory syncytial virus (major cause of the common cold) and influenza virus^15-17^. Monocytes undergo epigenetic reprogramming following exposure to Bacille Calmette-Guérin (BCG) vaccination, causing enhanced production of inflammatory cytokines including TNFα up to 3 months after vaccination, in response to unrelated stimuli^18^. Further studies are necessary to determine whether reprogramming of monocytes following COVID-19 infection may confer protection against subsequent infections (SARS-CoV-2 or unrelated).

In summary, we have identified features of circulating monocytes in convalescent COVID-19 patients that correspond to resolution of lung injury including restoration of cytokine production in response to stimuli, expression of migration-associated molecules and recovery of monocyte exhaustion. Focusing immune modulation strategies on monocytes and leucocyte migration following hospital discharge provides the potential for targeted therapeutic strategies in patients with persistent COVID-19 associated symptoms.

## Data Availability

All data needed to evaluate the conclusions in the paper are present in the paper or the Supplementary Materials.

## Acknowledgements

This report is independent research supported by the UK Coronavirus Immunology Consortium (UK-CIC), and the North West Lung Centre Charity and the NIHR Manchester Clinical Research Facility at Wythenshawe Hospital. We acknowledge the Manchester Allergy, Respiratory and Thoracic Surgery Biobank for supporting this project and thank the study participants for their contribution. The views expressed in this publication are those of the authors and not necessarily those of the NHS, the National Institute for Health Research or the Department of Health. Angela Simpson, Tim Felton Paul Dark and Tracy Hussell are supported by the NIHR Manchester Biomedical Research Centre. The authors would like to acknowledge the Manchester Allergy, Respiratory and Thoracic Surgery Biobank, the Northern Care Alliance Research Collection tissue bank and the North West Lung Centre Charity for supporting this project. In addition, we would like to thank the Immunology community within the Lydia Becker Institute of Immunology and Inflammation, the core flow cytometry facility at the University of Manchester, the Manchester COVID-19 Rapid Response Group and the study participants for their contribution. **Funding:** This work was supported by The Kennedy Trust for Rheumatology Research who provided a Rapid Response Award for costs associated with the laboratory analysis of the immune response in COVID-19 patients to JRG, The Wellcome Trust (TH, 202865/Z/16/Z; 106898/A/15/Z which helped support some CIRCO members), The Wellcome Trust/Royal Society (ERM, 206206/Z/17/Z), the Lister Institute (JEK) and BBSRC (JEK BB/M025977/1, TNS BB/S01103X/1), the Medical Research Council (LP MR/R00191X/1) and the NIHR Manchester Biomedical Research Centre (AGM). The Oxford, Leicester and Manchester NIHR BRC provided support for study design and sample collection.

---

**Extended data 1.**
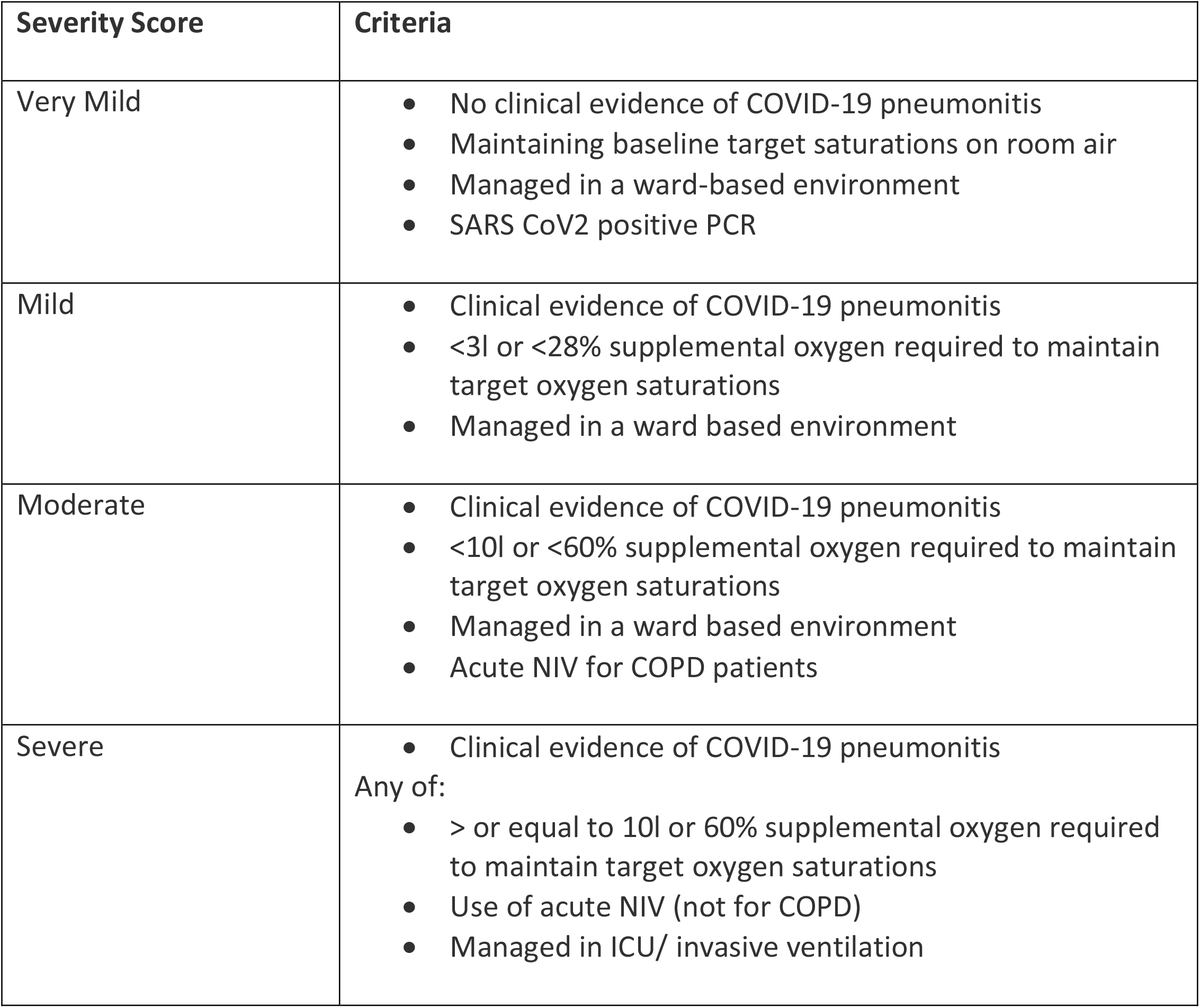
Patient categorization. Criteria for patient stratification. NIV: non-invasive ventilation; CPAP: continuous positive airway pressure; ICU: intensive care unit; COPD: chronic obstructive pulmonary disease.

**Extended data 2.**
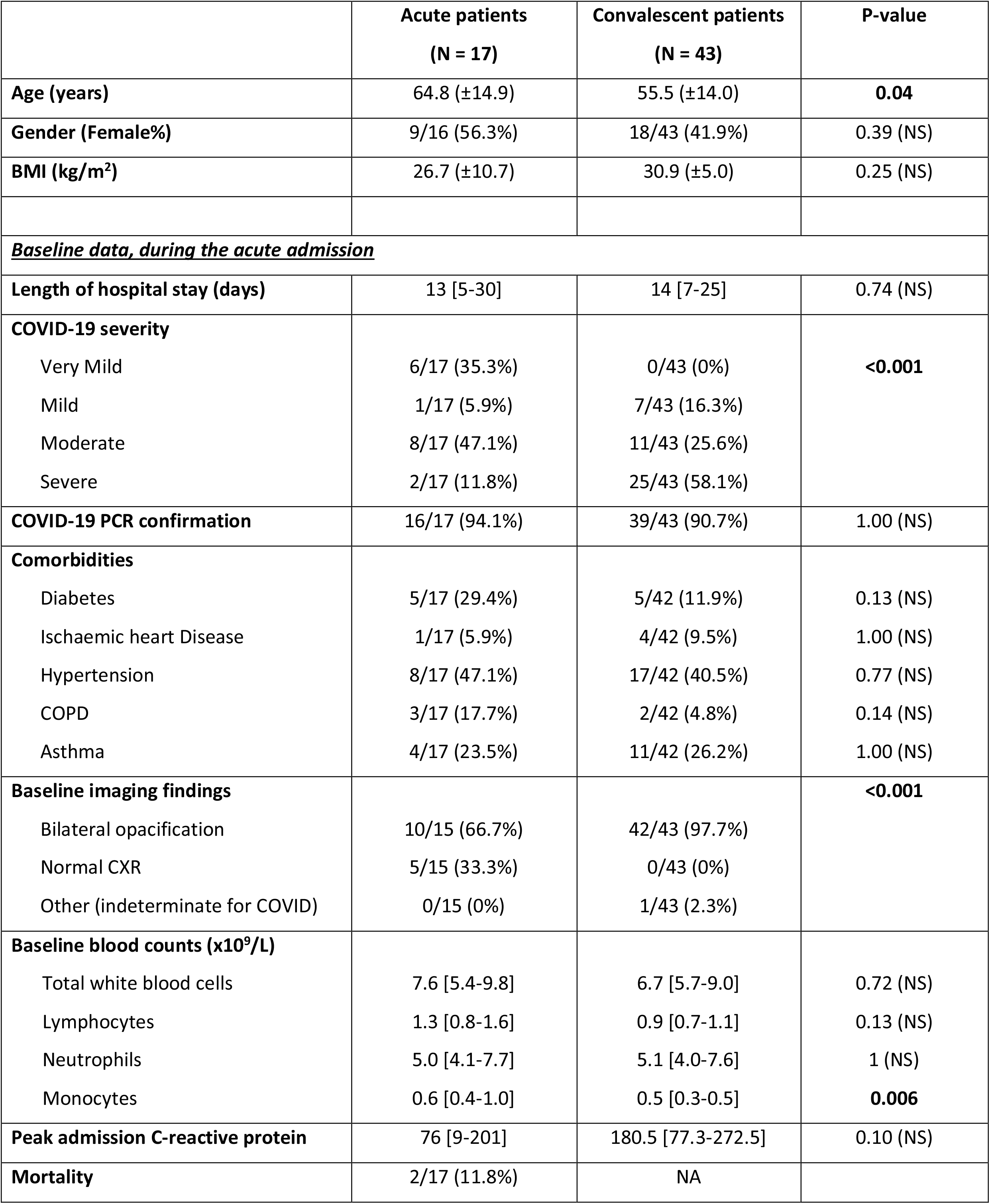
Clinical characteristics of acute versus convalescent patients. Normally distributed continuous data presented as mean (± standard deviation) and compared using t-test; while the remaining continuous data are presented as median [interquartile range] and compared using Mann-Whitney U test. Dichotomous data are presented as percentages and compared using chi-squared or the Fisher test, as appropriate.

**Extended data 3.**
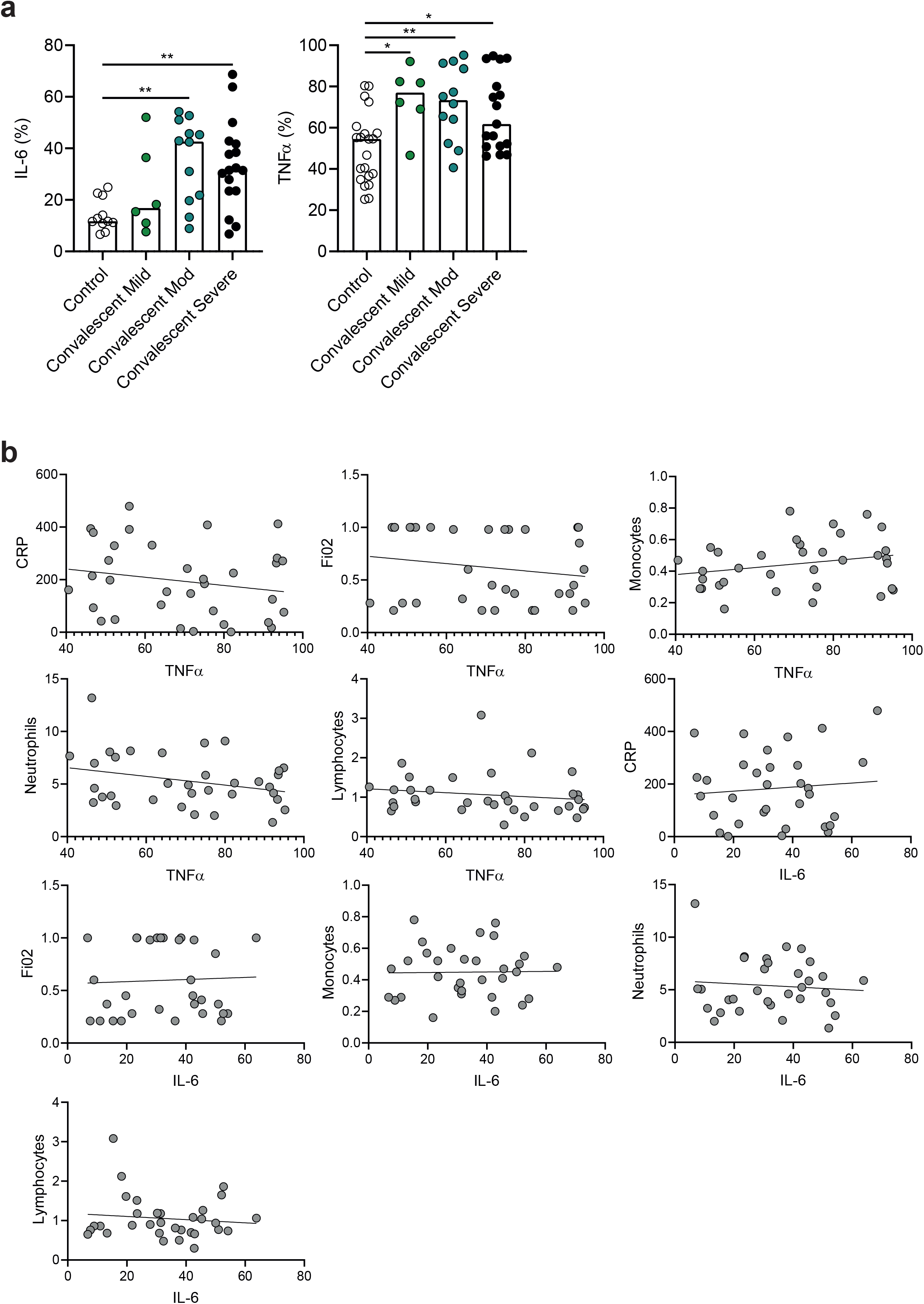
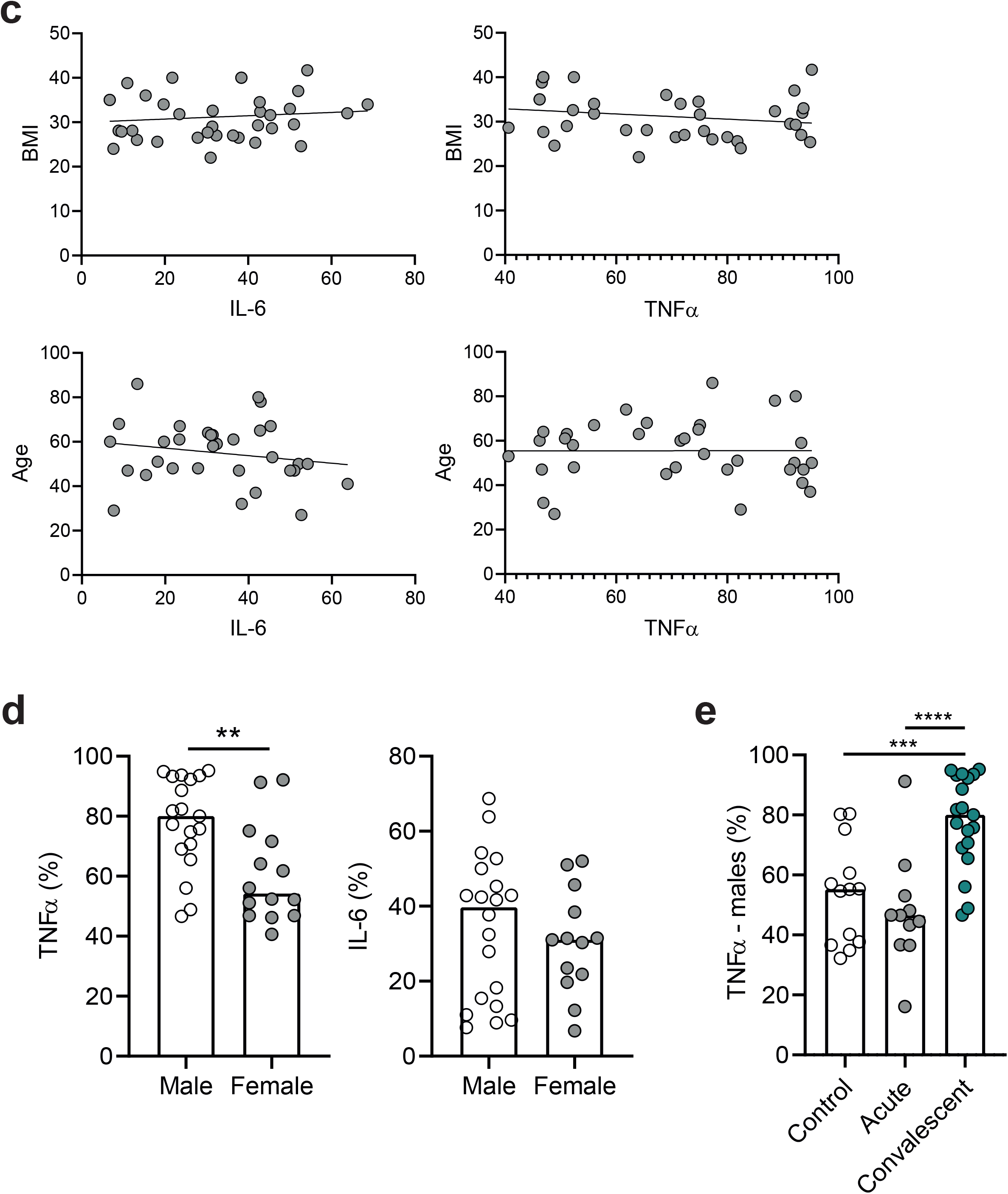
Correlations of cytokine production by monocytes during convalescence with clinical parameters during acute admission. **(A)** Graphs show proportions of monocytes expressing IL-6 (controls: n=11; mild: n=6; moderate: n=12; severe: n=17) and TNF*α* (controls: n=21; mild: n=6; moderate: n=12; severe: n=17) from convalescent COVID-19 patients, based on their disease severity during acute admission stage of disease. **(B)** Correlations of stimulated monocytes from convalescent COVID-19 patients producing TNF*α* versus CRP (n=34; Pearson correlation coefficient), FiO2 (n=34; Spearman’s ranked correlation coefficient), monocyte count (n=34; Pearson correlation coefficient), neutrophil count (n=34; Pearson correlation coefficient), lymphocyte count (n=34; Spearman’s ranked correlation coefficient); correlations of stimulated monocytes from convalescent COVID-19 patients producing IL-6 versus CRP (n=32; Pearson correlation coefficient), FiO2 (n=32; Spearman’s ranked correlation coefficient), monocyte count (n=32; Pearson correlation coefficient), neutrophil count (n=32; Pearson correlation coefficient), lymphocyte count (n=32; Spearman’s ranked correlation coefficient). **(C)** Correlations of BMI with monocyte production of IL-6 (n=36; Pearson correlation coefficient) and TNF*α* (n=37; Pearson correlation coefficient) in convalescence; correlations of age with monocyte production of IL-6 (n=32; Pearson correlation coefficient) and TNF*α* (n=34; Pearson correlation coefficient) in convalescence. **(D)** Graphs showing proportions of monocytes expressing TNF*α* (male: n=19; female: n=14) and IL-6 (male: n=20; female n=13) in male and female convalescent COVID-19 patients. **(E)** Graph showing proportions of monocytes expressing TNF*α* in male controls and male COVID-19 patients during acute admission or convalescence (control: n=13; acute: n=11; convalescent: n=19). One way ANOVA with Holm-Sidak post-hoc test: Ext data 3A and 3E; unpaired t-test: Ext data 3D. (P*<0.05; **P<0.01; ***P<0.001; ****P<0.0001).

**Extended data 4.**
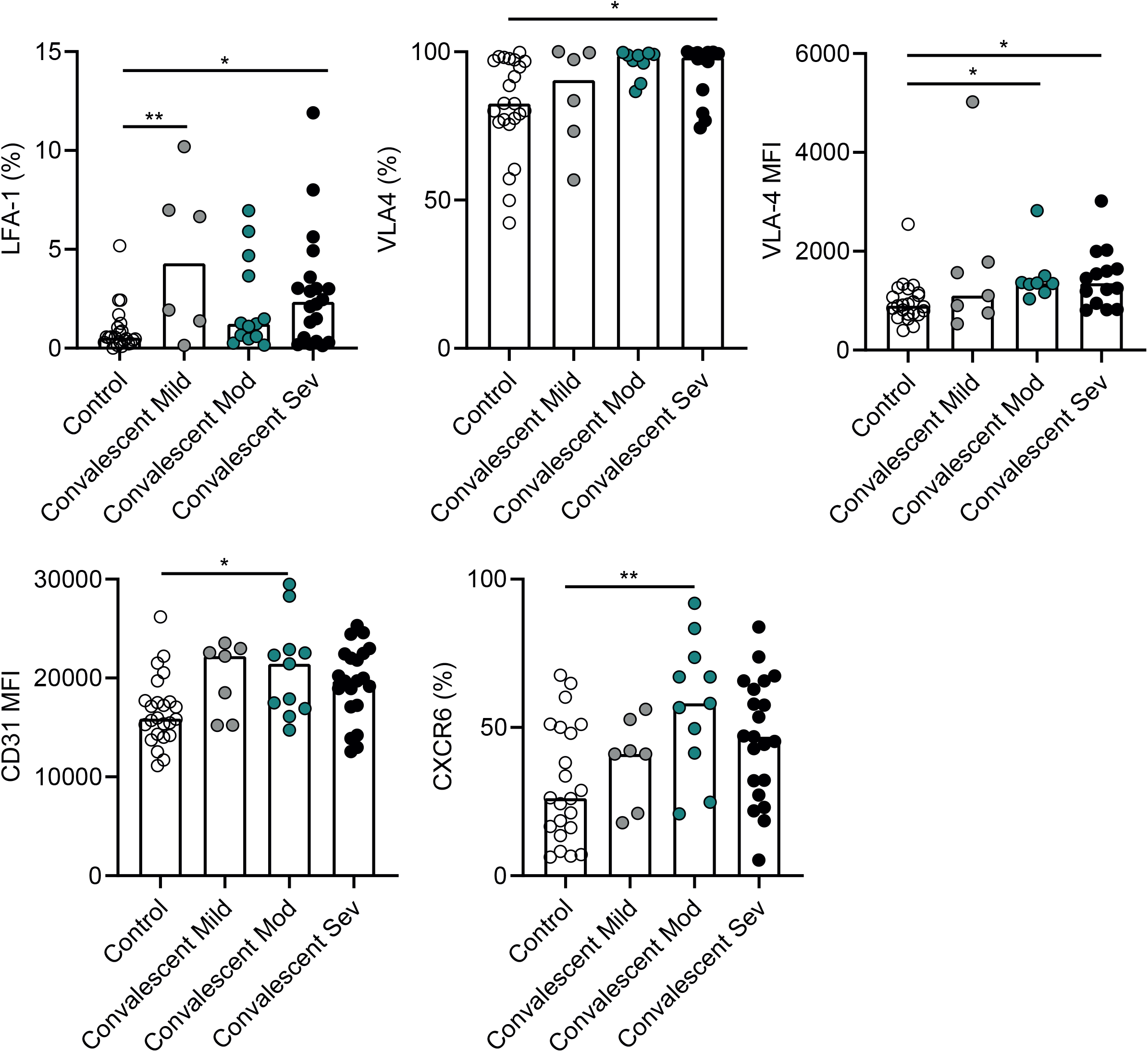
Correlations of migration marker expression by monocytes during convalescence with disease severity during acute admission. Graphs show proportions of monocytes expressing LFA-1 (controls: n=23; mild: n=6; moderate: n=13; severe: n=20), VLA-4 (controls: n= 23; mild: n=6; moderate: n=9; severe: n=14) and CXCR6 (controls: n=22; mild: n=7; moderate: n=11; severe: n=21), and levels of expression of VLA-4 (controls: n=23; mild: n=6; moderate: n=8; severe: n=14) and CD31 (controls: n=24; mild: n=7; moderate: n=11; severe: n=21) as assessed by MFI from convalescent COVID-19 patients, based on their disease severity during acute admission stage of disease. One way ANOVA with Holm-Sidak post-hoc test: LFA-1, VLA-4 %, CD31 MFI, CXCR6. Kruskal-Wallis test with Dunn’s post-hoc test: VLA-4 MFI. (P*<0.05; **P<0.01; ***P<0.001; ****P<0.0001).

**Extended data 5.**
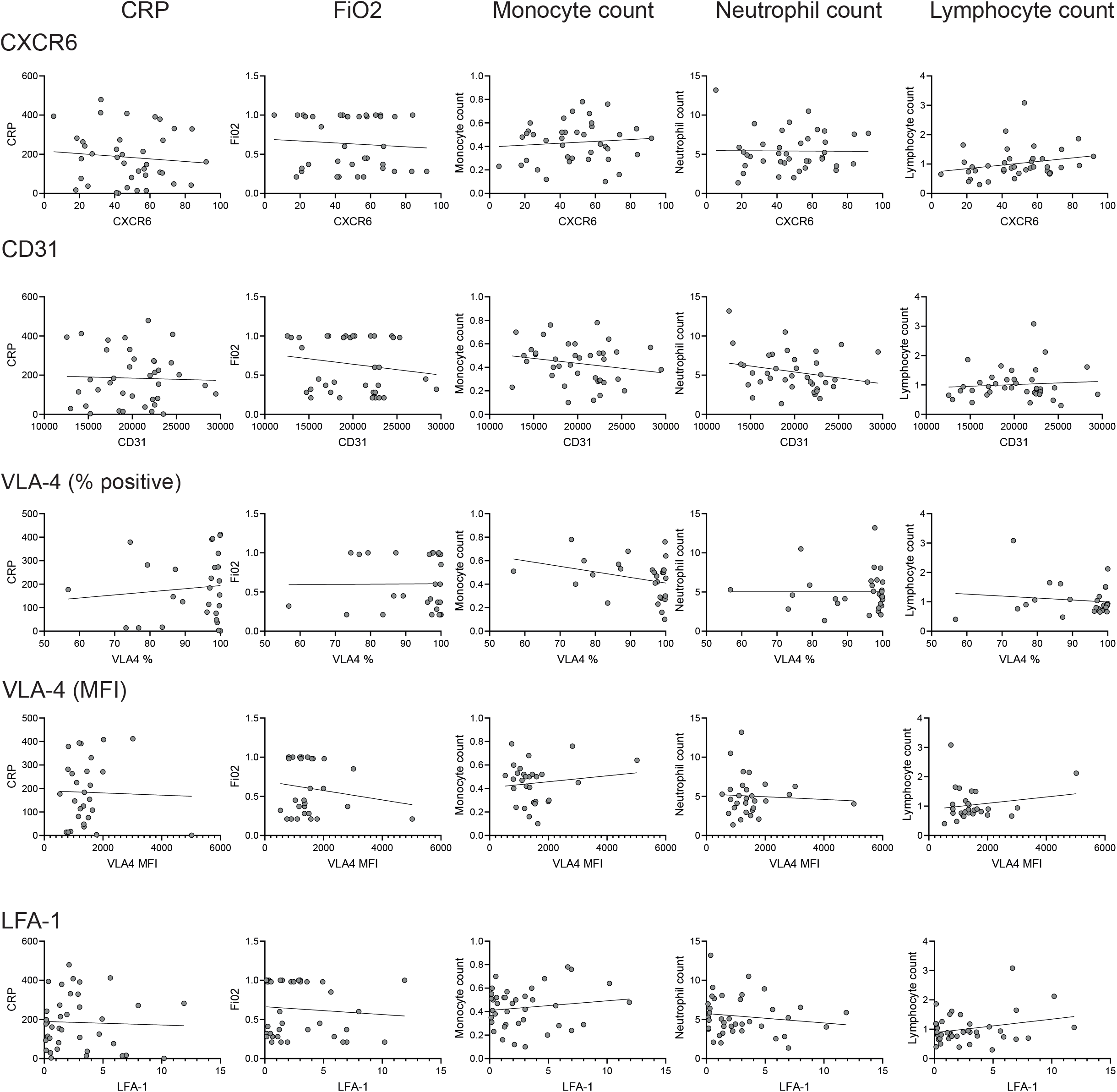
Correlations of migration marker expression by monocytes during convalescence with clinical parameters during acute admission. Correlations of monocytes from convalescent COVID-19 patients expressing CXCR6 with CRP (n=39; Pearson correlation coefficient), FiO2 (n=39; Spearman’s ranked correlation coefficient, monocyte count (n=40; Pearson correlation coefficient), neutrophil count (n=40; Pearson correlation coefficient) and lymphocyte count (n=40; Spearman’s ranked correlation coefficient). Correlations of monocytes from convalescent COVID-19 patients expressing CD31 with CRP (n=38; Pearson correlation coefficient), FiO2 (n=39; Spearman’s ranked correlation coefficient), monocyte count (n=39; Pearson correlation coefficient), neutrophil count (n=39; Pearson correlation coefficient), lymphocyte count (n=39; Spearman’s ranked correlation coefficient). Correlations of monocytes from convalescent COVID-19 patients expressing VLA-4 (% of monocytes) with CRP (n=28; Pearson correlation coefficient), FiO2 (n=29; Spearman’s ranked correlation coefficient), monocyte count (n=29; Pearson correlation coefficient), neutrophil count (n=29; Spearman’s ranked correlation coefficient) and lymphocyte count (n=29; Spearman’s ranked correlation coefficient). Correlations of levels of VLA-4 (MFI) on monocytes from convalescent COVID-19 patients with CRP (n=28; Pearson correlation coefficient), FiO2 (n=29; Spearman’s ranked correlation coefficient), monocyte count (n=29; Pearson correlation coefficient), neutrophil count (n=29; Spearman’s ranked correlation coefficient), lymphocyte count (n=29; Spearman’s ranked correlation coefficient). Correlations of monocytes from convalescent COVID-19 patients expressing LFA-1 with CRP (n=38; Pearson correlation coefficient), FiO2 (n=40; Spearman’s ranked correlation coefficient), monocyte count (n=40; Pearson correlation coefficient), neutrophil count (n=40; Pearson correlation coefficient) and lymphocyte count (n=40; Spearman’s ranked correlation coefficient).

**Extended data 6.**
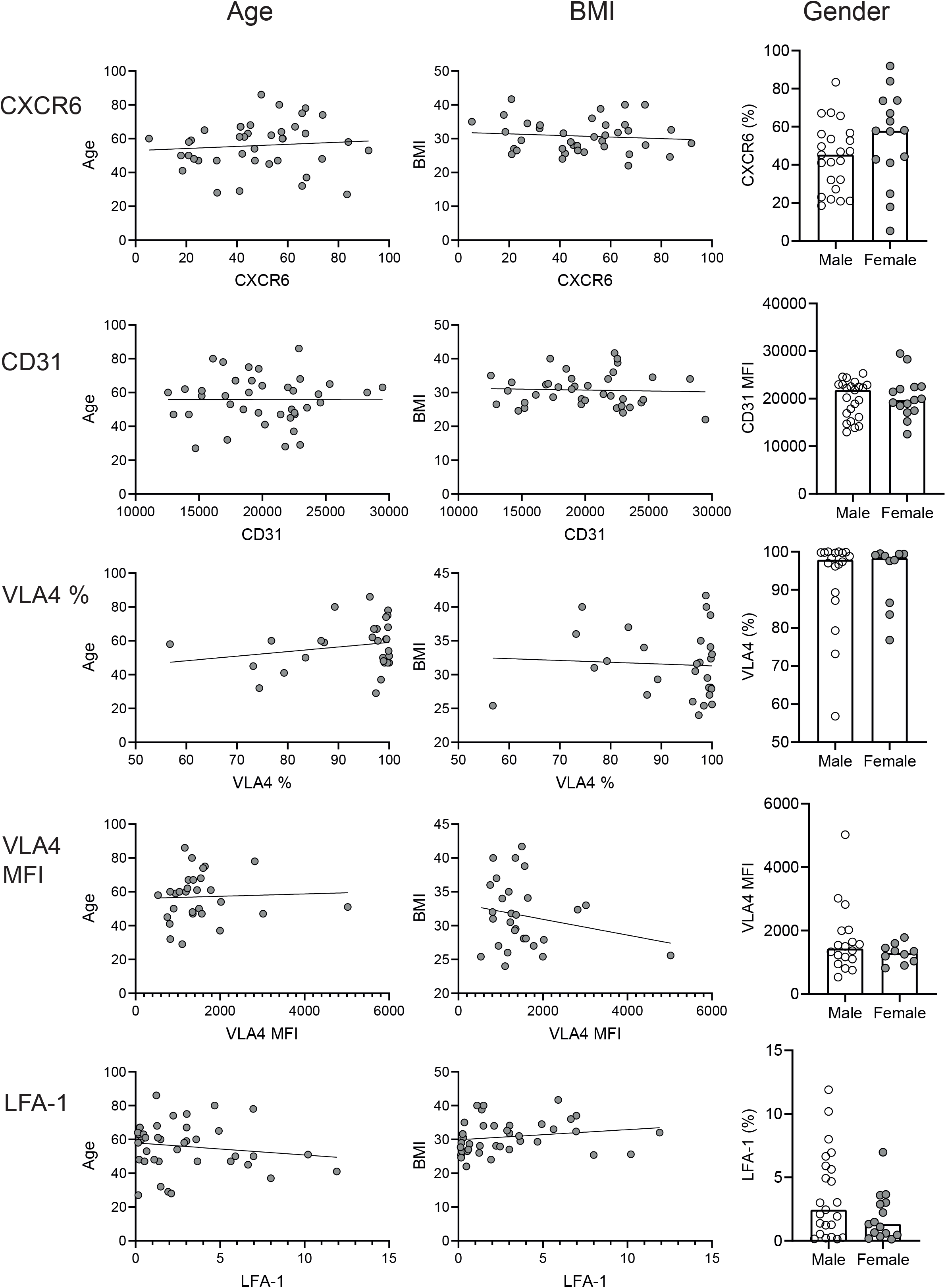
Correlations of migration marker expression by monocytes during convalescence with age, BMI and gender. Correlations of monocytes from convalescent COVID-19 patients expressing CXCR6 with age (n=40; Spearman’s ranked correlation coefficient) and BMI (n=39; Pearson correlation coefficient) and graph showing proportions of monocytes expressing CXCR6 in male (n=23) and female (n=15) convalescent patients. Correlations of CD31 levels (MFI) on monocytes from convalescent COVID-19 patients with age (n=40; Pearson correlation coefficient) and BMI (n=38; Pearson correlation coefficient) and graph showing levels of CD31 (MFI) in male (n=23) and female (n=15) convalescent patients. Correlations of monocytes from convalescent COVID-19 patients expressing VLA-4 with age (n=29; Pearson correlation coefficient) and BMI (n=28; Pearson correlation coefficient) and graph showing proportions of monocytes expressing VLA-4 in male (n=18) and female (n=10) convalescent patients. Correlations of VLA-4 levels (MFI) on monocytes from convalescent COVID-19 patients with age (n=29; Pearson correlation coefficient) and BMI (n=28; Pearson correlation coefficient) and graph showing levels of VLA-4 (MFI) on monocytes in male (n=18) and female (n=10) convalescent patients. Correlations of monocytes from convalescent COVID-19 patients expressing LFA-1 with age (n=40; Pearson correlation coefficient) and BMI (n=39; Pearson correlation coefficient) and graph showing proportions of monocytes expressing LFA-1 in male (n=23) and female (n=15) convalescent patients. Unpaired t-test: CXCR6, CD31 MFI, VLA-4 %, VLA-4 MFI. Unpaired Mann-Whitney test: LFA-1. (P*<0.05; **P<0.01; ***P<0.001; ****P<0.0001).

