## Supplemental methods for "Recovery of monocyte exhaustion is associated with resolution of lung injury in COVID-19 convalescence"

### Study design

Between 14th June and 25th August, 2020, 44 patients attending outpatient follow up 8-19 weeks after discharge following inpatient admission for COVID-19 were included as “convalescent” patients. Two comparator groups were included in analysis; “acute” patients recruited during in-patient stays for COVID-19 and healthy controls. Twenty “acute” patients with inpatient admissions between 14th June and 12th August 2020 were included. Informed consent was obtained for each patient. Peripheral blood samples were collected at Manchester University Foundation Trust (MFT), Salford Royal NHS Foundation Trust (SRFT) and Pennine Acute NHS Trust (PAT) under the framework of the Manchester Allergy, Respiratory and Thoracic Surgery (ManARTS) Biobank (study no M2020-88) for MFT or the Northern Care Alliance Research Collection (NCARC) tissue biobank (study no. NCA-009) for SRFT and PAT (REC reference 15/NW/0409 for ManARTS and 18/WA/0368 for NCARC). Clinical information was extracted from written/electronic medical records. Patients were included if they tested positive for SARS-CoV-2 by reverse-transcriptase–polymerase-chain-reaction (RT-PCR) on nasopharyngeal/oropharyngeal swabs or sputum, during acute admission. Patients with negative nasopharyngeal RT-PCR results were also included if there was a high clinical suspicion of COVID-19, the radiological findings supported the diagnosis and there was no other explanation for symptoms, upon acute admission. Patients were excluded if an alternative diagnosis was reached, or where indeterminate imaging findings were combined with negative SARS-CoV-2 nasopharyngeal (NP) test. The severity of disease was scored each day during acute admission, based on degree of respiratory failure (**Extended Data 1**). Where severity of disease changed during admission, the highest disease severity score was selected for classification. Peripheral blood samples were collected as soon after admission as possible. Healthy blood samples were obtained from frontline workers at Manchester University and NHS Trusts (age range 28-69). Samples from healthy donors were examined alongside patient samples.

**Isolation of PBMCs**

Whole venous blood was collected in tubes containing EDTA or serum gel clotting activator (Starstedt). Peripheral blood mononuclear cells (PBMCs) were isolated by density gradient centrifugation using Ficoll-Paque Plus (GE Healthcare) and 50 ml SepMate tubes (STEMCELL technologies) according to the manufacturer's protocol. Serum was separated by centrifuging serum tubes at 2000 × g at 4°C for 20 min.

**Flow cytometry**

White blood cells from lysed whole blood and isolated PBMCs separated by density gradient centrifugation were stained immediately on receipt. The following antibodies were used, CCR7 (clone G043H7), CD14 (clone 63D3), CD16 (clone 3G8), CD19 (clone H1B19), CD3 (clone OKT3 or UCHT1), CD45 (clone 2D1), HLA-DR (clone L234), $K_i$-67 (clone $K_i$-67 or 11F6), CD66b (clone G10F5), CD64 (clone 10.1), IL-1β (clone H1b-98), TNF-α (clone MAb11), CCL2 (clone 5D3-F7), IL-6 (clone MQ2-13A5), LFA-1 (clone M24), CD31 (clone WM59) all from Biolegend; and COX-2 (clone AS67), CXCR6 (clone 13B 1E5), CD62L (clone SK11) and VLA-4/CD49d (clone L25) from BD Biosciences.

PBMCs were also stimulated in vitro for 18-20 hours with 10 ng/ml LPS in the presence of 10 μg/ml brefeldin A to allow accumulation and analysis of intracellular proteins by flow cytometry. Cells were cultured in RPMI containing 10% fetal calf serum, L-Glutamine, Non-essential Amino Acids, HEPES and penicillin plus streptomycin (Gibco). For surface stains samples were fixed with BD Cytofix (BD Biosciences) prior to acquisition and for intracellular stains ($K_i$-67, COX-2, TNF-α and IL-1β) the Foxp3/Transcription Factor Staining Buffer Set (eBioscience) was used. All samples were acquired on a LSRFortessa flow cytometer (BD Biosciences) and analyzed using FlowJo (TreeStar).

**Statistics**

Results are presented as individual data points with medians. Statistical analysis was performed using Prism 8 Software (GraphPad). Normality tests were performed on all datasets. Groups were compared using an unpaired *t*-test (normal distribution) or Mann-Whitney test (failing normality testing) for healthy individuals versus COVID-19 patients. One-way ANOVA with Holm-Sidak post-hoc testing (normal distribution) or Kruskal-Wallis test with Dunn's post-hoc testing (failing normality testing) was used for multiple group comparisons. Correlations were assessed with Pearson correlation coefficient (normal distribution) or Spearman's rank correlation coefficient test (failing normality testing) for separate parameters within the COVID-19 patient group. Information on tests used is detailed in figure legends. In all cases, a *p*-value of ≤ 0.05 was considered significant. ns, not significant; *$p < 0.05$, **$p < 0.01$, ***$p < 0.001$.
